# Altered white matter microstructure of language pathways and semantic cognition deficiencies in early psychosis

**DOI:** 10.1101/2025.05.29.25328555

**Authors:** Werner Surbeck, Wolfgang Omlor, Noemi Dannecker, Robin Samuel, Anna Steiner, Dominic Fabian, Allegra Gasser, Roya Hüppi, Rahel Horisberger, Giacomo Cecere, Nils Kallen, Pierfrancesco Sarti, Philipp Stämpfli, Felix Scholtes, Niklaus Denier, Tobias Bracht, Peter Brugger, Philipp Homan

## Abstract

**Background:** Semantic language dysfunction is a hallmark of early psychosis, yet the underlying brain structural correlates are largely unexplored. In particular, it is unclear whether core deficits arise from disruptions to semantic representation, which refers to the stored knowledge of word meanings, or to semantic control, which entails top-down mechanisms that guide the retrieval and selection of context-appropriate semantic information. By dissociating semantic representation from semantic control, we aimed to clarify which aspect of semantic processing is preferentially impaired in early psychosis and how these deficits map onto structural properties of the ventral and dorsal language streams.

**Methods:** We investigated N = 120 individuals across the psychosis spectrum: N = 40 individuals with early psychosis, N = 40 individuals with high schizotypy and N = 40 individuals with low schizotypy. Participants with high and low schizotypy constituted the non-clinical comparison group. All participants completed tasks designed to isolate semantic representation and semantic control processes. Given the importance of accurate delineation, this study employed meticulous manual fiber tractography of diffusion tensor imaging (DTI) data to ensure reliable evaluation of ventral and dorsal pathway microstructure.

**Findings:** Compared to individuals with high and low schizotypy, individuals with early psychosis showed pronounced deficits in semantic control while semantic representation remained largely intact. Mean diffusivity in the left inferior fronto-occipital fasciculus and left uncinate fasciculus was lower in the early psychosis group than in individuals with schizotypy. In the early psychosis group, fractional anisotropy in the left arcuate fasciculus was negatively correlated with semantic control but no DTI measure was associated with semantic representation.

**Conclusion:** These results underscore semantic control as a core deficit in early psychosis and extend the conventional view that semantic processing is subserved primarily by ventral pathways. The arcuate fasciculus appears implicated in semantic control, indicating a more integrated interplay of dorsal and ventral streams in semantic language processing.

## INTRODUCTION

Semantic processing abnormalities, clinically manifested as formal thought disorder and delusional phenomena, are considered core symptoms of schizophrenia ^1,2^. Similar, albeit milder, semantic disruptions have been described in individuals with schizotypy—a cluster of personality traits thought to arise from a similar combination of genetic, neurodevelopmental, and psychosocial factors as schizophrenia, placing schizotypy on a continuum with the disorder ^3^.

Evidence suggests that white matter pathway organization is linked to distinct symptom domains in schizophrenia ^4^. Recent research indicates that formal thought disorder and delusions in schizophrenia correlate with altered white matter microstructure in the left dorsal language stream as well as in the left ventral language stream ^5–8^. Moreover, altered microstructure of ventral language stream pathways has been linked to subclinical cognitive-perceptual (positive) symptoms of schizotypy in a multimodal lesion-mapping study ^9^. Direct electrical stimulation of the inferior fronto-occipital fasciculus (IFOF)—a principal component of the ventral language stream—elicits semantic paraphasias when applied on the left side during awake surgery and impairs nonverbal semantic association tasks bilaterally ^10–12^. These observations underscore the essential role of the IFOF in semantic processing and suggest that alterations in this pathway could contribute to semantic anomalies in schizophrenia-spectrum disorders. Consistent with this idea, white matter microstructure of the ventral language stream, particularly the left IFOF, has been shown to be associated with semantic processing deficits in schizophrenia ^13^.

However, deficits in semantic processing may arise from disruptions in two complementary facets of meaning processing: semantic representation and semantic control ^14^. Semantic representation refers to stored knowledge about words and concepts, encompassing the fundamental meanings that accumulate over a lifetime ^15^. In contrast, semantic control involves the goal-directed retrieval, selection, and integration of relevant semantic information from these stores, while inhibiting competing or irrelevant associations ^14^. For example, whereas an intact semantic representation allows an individual to know that a “palm” can refer to both a part of the hand and a type of tree, semantic control determines which meaning is contextually appropriate when the individual hears the word “palm” in conversation. While insights into these distinct components have emerged from neurology and neurosurgery ^16^, their precise structural correlates remain unclear—particularly within schizophrenia-spectrum disorders. The white matter pathways that support semantic representation and control are principally located within the ventral language stream, which includes both an indirect route – comprising the inferior longitudinal fasciculus (ILF) and the uncinate fasciculus (UF), and a direct route via the IFOF ^17^.

The present study aims to clarify how perisylvian white matter microstructure relates to specific dimensions of semantic cognition—namely, semantic representation and semantic control—in individuals with early psychosis. We hypothesized that deficits in semantic representation and control are associated with altered white matter microstructure of ventral language stream components, thereby illuminating the neural characteristics of language disturbances commonly observed in the early stages of schizophrenia-spectrum disorders.

## MATERIAL AND METHODS

### Subjects

As part of the VELAS (Ventral language stream in schizophrenia with regard to semantic and visuo-spatial processing anomalies) study ^18–20^, 12 female and 28 male (total N = 40) early psychosis patients and 41 female and 39 male (total N = 80) individuals with high (N = 40) and low (N = 40) schizotypy, matched for age and education, were included in this study. Subjects were recruited from the inpatient and outpatient departments of the Psychiatric Hospital of the University of Zurich, Switzerland. Patients fulfilled the International Statistical Classification of Diseases and Related Health Problems 10th Revision (ICD-10) criteria for a psychotic disorder: schizophrenia (F20; N = 22), acute and transient psychotic disorder (F23; N = 10), schizoaffective disorder (F25; N = 5), recurrent depressive disorder or current episode severe with psychotic symptoms (F33.3; N = 3). Individuals with schizotypy were recruited through an online pre-assessment, which was spread over different platforms and noticeboards of Swiss universities and colleges, common Swiss public platforms, a mailing list of students of psychology, tweets by the research team and direct contact of different schools and educational institutions of various levels. For the pre-assessment, an online schizotypy screening (N *=* 1062) was conducted using the Oxford-Liverpool Inventory of Feelings and Experiences (O-LIFE) ^21^ and Multidimensional Schizotypy Scale (MSS) ^22^. Using model-based clustering (mclust in R) according to Scrucca, et al. ^23^, we identified five (MSS) and four (O-LIFE) schizotypy clusters. Using the predict.Mclust function ^23^, we determined which cluster each of the 40 patients would most likely belong to based on their schizotypy scores. This showed that all patients would most likely belong to MSS clusters 3L5 and O-LIFE clusters 2L4, i.e. clusters with higher schizotypy scores. 40 healthy controls from the same clusters, i.e. matching the patients in schizotypy (in addition to age and education), were selected for the “high schizotypy” group. 40 healthy controls from MSS clusters 1L2 and O-LIFE cluster 1, i.e. with lower schizotypy scores (matching the patients in age and education) were selected for the “low schizotypy” group. To be included, all subjects had to be aged 14 – 40 years and right-handed as determined by the short form of the Edinburgh handedness inventory ^24^. Participants with a current substance use disorder (excluding nicotine) were excluded. Exclusion criteria also included past or current neurological or ophthalmological disorders, history of head trauma with concurrent loss of consciousness, and current pregnancy. Exclusion criteria limited to control subjects were a history of any psychiatric disorder. All but 6 patients were under antipsychotic pharmacotherapy. All participants provided written informed consent, the ethics committee of Kantonale Ethikkommission Zürich gave ethical approval for this work (KEK-ZH 2020/01049) and the study adhered to the Declaration of Helsinki.

### Behavioral data acquisition

#### Symptom assessments

Assessment of schizophrenia psychopathology was conducted on the day of testing and included the Positive and Negative Syndrome Scale (PANSS) ^25^ as well as the O-LIFE ^21^ and MSS ^22^ in all participants.

#### Neuropsychological assessments

Participants completed a picture naming task (DO80) to assess neuropsycholinguistic impairment ^26^, as well as the multiple-choice word test (MWT-B) to evaluate semantic representation ^27^, verbal fluency tasks ^28,29^ and the Camel and Cactus Test (CCT), which requires matching an image with a matching image from a selection of four images based on their semantic association, to measure semantic control ^30^. Visual stimuli for the CCT were generated using custom scripts based on the PsychoPy software suite ^31^.

Finally, we controlled for effects of executive functions, processing speed and attention, which have been shown to be relevant for semantic processing ^32^. These were assessed using the Victoria Stroop ^33^, the Trail Making Test (TMT) A and B ^34^, as well as the digit span forward and backward ^35^.

#### MRI data acquisition

Imaging was performed on a Philips Achieva 3.0 T magnetic resonance scanner with a 32 channel SENSE head coil (Philips, Best, The Netherlands). 3D-T1-weighted images with a voxel resolution of 1mm^3^ as well as diffusion weighted imaging (DWI) with 64 non-collinear directions (3000 s/mm^2^) and one b0 (0 s/mm^2^), were acquired with a voxel resolution of 1.96×1.96×2mm^3^.

#### Diffusion tensor imaging

We used DWI to perform diffusion tensor imaging (DTI). First, we used HD-BET, a high-quality deep learning based algorithm, for brain extraction using the T1-weighted images ^36^. Second, for the computation of the diffusion tensor out of DWI, we used ExploreDTI 4.8.6 ^37^. We applied a subject motion and distortion correction to DWI data (reference volume: b0 image). Second, we warped the resulting data to the brain-extracted T1 volumes using an EPI correction ^38^. We performed whole-brain deterministic tractography with a diffusion tensor model ^39^. Tract termination criteria were set to fractional anisotropy < 0.2 and tract angle > 45 degrees.

#### Manual tractography

For the segmentation of specific bilateral tracts, manual tractography was performed. We used region-of-interest (ROI) masks for the selection of specific fibers of the whole brain DTI dataset.

To reconstruct the arcuate fasciculus (AF) (**Fig. 2a**), the first ROI was positioned around the deep white matter at the fronto-parietal junction, where the AF appears as a green layer in the coronal plane of the directionally encoded tensor (DET) map, situated laterally to the blue fibers of the corona radiata. The second ROI is placed around the descending segment of the superior longitudinal fasciculus in the posterior temporal lobe, visible as a blue structure lateral to the splenium of the corpus callosum in the transverse plane of the DET map ^40^.

To track the IFOF (**Fig. 2b**), the first ROI is placed around the entire frontal lobe on a coronal slice at the level of the precentral sulcus. The second coronal ROI is positioned to encompass the entire hemisphere at the junction of the parieto-occipital and calcarine sulci, identified on the mid-coronal slice, following the approach of Smits, et al. ^40^ with slight modifications.

The UF (**Fig. 2d**) is tracked by selecting the most posterior coronal slice where the temporal lobe is distinct from the frontal lobe. The first ROI encompasses the entire temporal lobe, while the second ROI includes the full set of projections toward the frontal lobe, appearing green on the DET map ^41^.

To track the ILF (**Fig. 2c**), the first ROI is positioned within the white matter of the anterior temporal lobe, just anterior to the temporal horn of the ventricular system. The second ROI is placed in the coronal plane at the level of the preoccipital notch ^42^.

To track the corticospinal tract (CST), the first transversal ROI was placed at the level of the pons, with the identification of CST (**Fig. 2e**) fibers facilitated by the DET map, where the pyramidal tract appears blue. The second ROI was positioned in the axial plane around the precentral gyrus, at the base of the precentral and central sulci ^43^.

After generating the fiber tracts based on the described ROIs, we manually removed fibers that belonged to neighboring fiber systems using carefully placed regions of avoidance. Finally, mean values of fractional anisotropy, mean diffusivity, axial diffusivity and radial diffusivity were extracted from reconstructed tracts using in-house software.

#### Statistical analysis

Group comparisons of white matter fiber pathways were performed using ordinary least squares regression models, adjusted for age, sex, education, as well as attention and executive functions. Coefficients within models were compared using Wald tests, and coefficients between models were compared using seemingly unrelated estimation adjusted as above. Group comparisons of semantic processing were conducted using random-effects multinomial logistic regression models, adjusted as above, reporting relative-risk ratios (RRR). Response latencies were assessed using multilevel mixed-effects linear regression models with a clustered sandwich estimator, adjusted as above. The association between white matter microstructure and impairments in semantic processing was analyzed using negative binomial regression models, reporting incidence-rate ratios (IRR) and random-effects multinomial logistic regression models, as appropriate and as adjusted as above. Coefficients between models were compared using seemingly unrelated estimation adjusted as above. All tests were performed using Stata version 18 (StatCorp, Texas). Estimates were considered statistically significant when *p* < .05. Unless otherwise stated, two-tailed hypothesis testing was used and unadjusted p values were reported.

## RESULTS

### Demographic and global brain measures

**Table 1** summarizes the demographic characteristics and clinical data for individuals with psychosis as well as for individuals with low and high schizotypy. Compared to controls, the patient group included a higher percentage of males, although this difference was not statistically significant (chi2 = 5.34, *p* = 0.07). There was also no significant difference in age between the groups (ANOVA: *F* = 2.94, *p* = 0.06). Behavioral data for different language tasks is presented in **Supplementary Table 1**.

**Table 1:** Sample Characteristics.

|  | Low schizotypy<br>(*n* = 40) | High schizotypy<br>(*n* = 40) | Patient (*n* = 40) |
| --- | --- | --- | --- |
| Sex, *n* (%) |  |  |  |
| Female | 22 (55) | 19 (47.5) | 12 (30) |
| Male | 18 (45) | 21 (52.5) | 28 (70) |
| Age [years], *M* (*SD*) | 25.1 (3.8) | 27.5 (4.6) | 27.2 (5.7) |
| Education, *n* |  |  |  |
| Secondary School | 0 | 0 | 8 |
| Vocational Training | 11 | 10 | 17 |
| Higher Vocational Training | 3 | 2 | 0 |
| Baccalaureate ("Matura") | 11 | 9 | 3 |
| University (Bachelor's, incl.<br>Applied Sciences) | 11 | 12 | 5 |
| University (Master's / Doctorate) | 4 | 7 | 7 |
| First language, *n* (%) |  |  |  |
| German | 31 (77.5) | □ | 7 (17.5) |
| Bilingual | 8 (20) | □ | □ |
| Other | 1 (2.5) | □ | 31 (77.5) |
| Diagnosis, *n* (%) |  |  |  |
| Schizophrenia (F20) | □ | □ | 22 (55) |
| Acute and transient psychotic<br>disorder (F23) | □ | □ | 10 (25) |
| Schizoaffective disorder (F25) | □ | □ | 5 (12.5) |
| Recurrent depressive disorder<br>with severe psychotic symptoms<br>(F33.3) | □ | □ | 3 (7.5) |
| Age onset [years], *M* (*SD*) | □ | □ | 26.2 (7.1) |
| Duration of illness [weeks], *M* (*SD*) | □ | □ | 128.6 (119.7) |
| No. of episodes, *n* (%) |  |  |  |
| 1 | □ | □ | 22 (55) |
| 2 | □ | □ | 12 (30) |
| 3 | □ | □ | 5 (12.5) |
| 4 | □ | □ | 1 (2.5) |
| Type of antipsychotic therapy, *n* (%) |  |  |  |
| None | 40 (100) | 40 (100) | 6 (15) |
| Current antipsychotic<br>monotherapy | □ | □ | 27 (67.5) |
| Current intake of 2 antipsychotics | □ | □ | 7 (17.5) |
| Duration of antipsychotic medication<br>[weeks], *M* (*SD*) | □ | □ | 56.9 (88.5) |

### Group comparisons of semantic processing

Individuals with early psychosis showed significantly more semantic and phonematic paraphasias in the DO80 than both schizotypy controls (RRR 1.419, 95% CI [1.143, 1.762], p = 0.002 and 4.448, [1.523, 12.990], p = 0.006, respectively), as calculated with random-effects multinomial logistic regression models. With regard to semantic representation, there was no difference between individuals with early psychosis and with schizotypy (**Fig. 1**). By contrast, there were significant differences with regard to semantic control, where patients with early psychosis named significantly fewer items within one minute than schizotypy controls in the domains clothing and food (clothing: IRR: 0.832, 95% CI: 0.75, 0.922, p < 0.001; food: IRR: 0.899, 95% CI: 0.815, 0.991, p = 0.03) but not in the domain animals (**Fig. 1**). In the CCT, patients showed a significantly longer response latency than schizotypy controls (1.295, 95% CI [0.682, 1.908], p < 0.001). This increased response latency was more pronounced compared with controls with low schizotypy (1.517, 95% CI [0.882, 2.152], p < 0.001).

**Figure 1:**
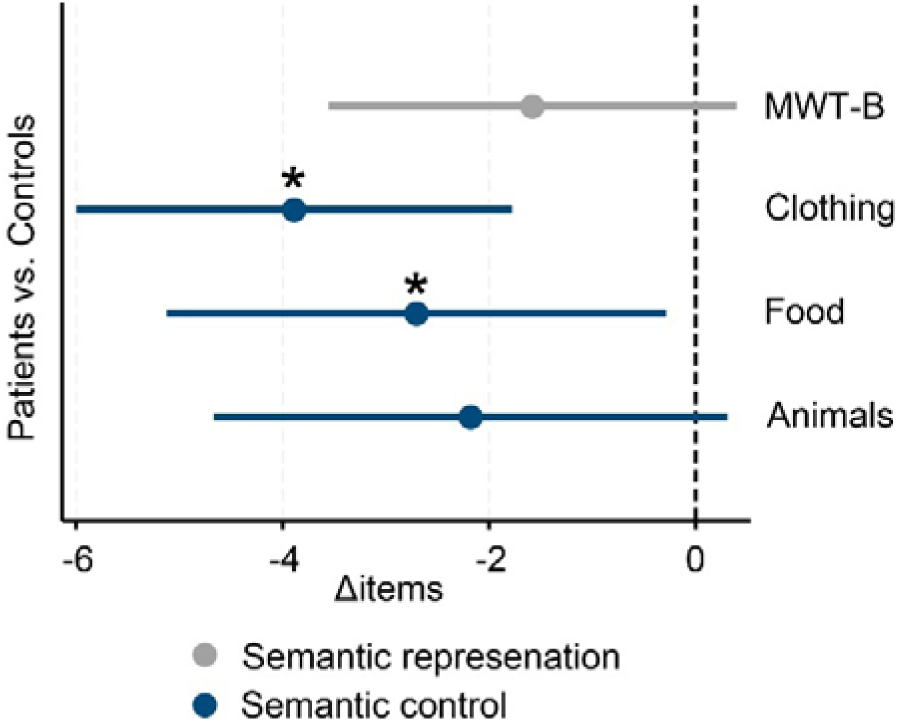
Group differences in semantic representation and control between individuals with early psychosis and schizotypy. Semantic representation did not differ significantly between groups. However, individuals with early psychosis exhibited significantly reduced semantic control compared to schizotypy controls, as reflected in fewer items named within one minute in the clothing and food domains, but not in the animal domain. Differences shown are average marginal effects based on estimates from negative binomial regression models reported above. Error bars represent 95% confidence intervals. Asterisks indicate p < 0.05.

### Group comparisons of white matter fiber pathways

DTI metrics on ventral and dorsal language streams are shown in **Supplementary figures 1 and 2** and language-related white matter pathways investigated with manual fiber tractography are illustrated in **Fig. 2**. With regard to the ventral language stream, early psychosis patients had lower mean diffusivity than controls with regard to the left IFOF (−0.486 standard deviation, 95% CI [−0.913, −0.058], t = −2.251, p = 0.026) and the left UF (−0.796 standard deviation, 95% CI [−1.199, −0.392], t = −3.909, p < .001) as calculated by regression models of the mean diffusivity adjusted for age, gender, education, attention, and executive functions. The difference between early psychosis and high schizotypy was significantly greater than between early psychosis and low schizotypy for the left UF (F = 4.895, p = 0.029) as established by a Wald test. Although no significant results were found in relation to the left AF per se, there were no significant differences of the mean diffusivity measures between the IFOF and the AF. In contrast, there was a significant difference in mean diffusivity measures between the UF and the AF (chi2(1) = 4.15, p = 0.042), as found with seemingly unrelated estimation adjusted as above. No significant group differences were found with regard to the CST.

**Figure 2.**
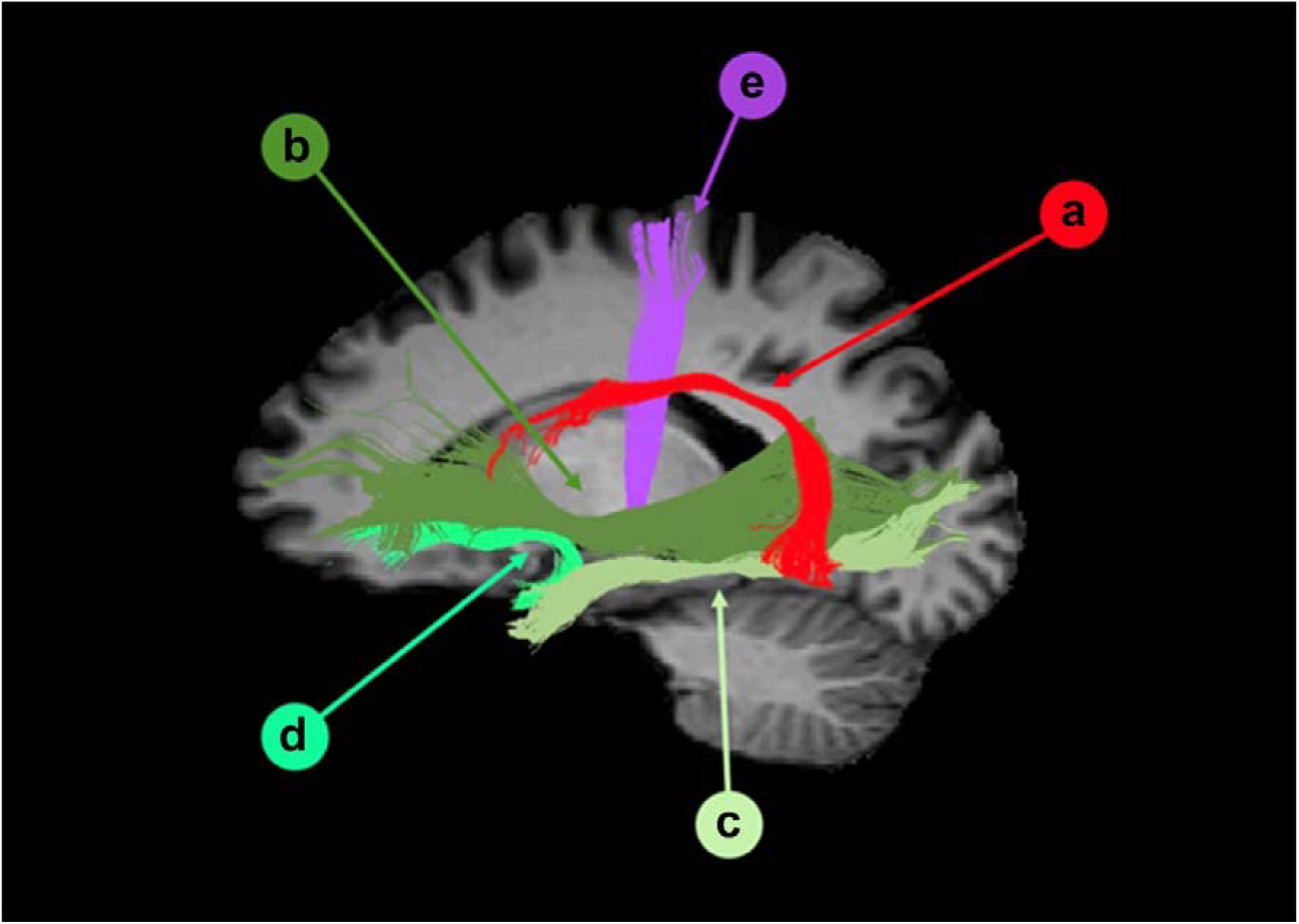
DTI-based manual fiber tractography of language-related white matter pathways. Results from one representative individual with early psychosis is illustrated (left lateral view). Language processing mainly relies on a dual-stream architecture: a dorsal stream – including the arcuate fasciculus (a) – and a ventral stream. The latter depends on a direct pathway, the inferior fronto-occipital fasciculus (b), and an indirect pathway, formed by the inferior longitudinal fasciculus (c) and uncinate fasciculus (d). The corticospinal tract (e) is also shown.

### Association of tract microstructure with impairments in semantic processing

Diffusion measures fractional anisotropy and mean diffusivity of the tracts examined showed no significant (p < 0.05) association with semantic representation either within the patient group or within the control group. In contrast, fractional anisotropy measures were negatively associated with verbal fluency as measured with naming items within one minute with regard to the left AF (clothing: IRR 0.911, 95% CI [0.842, 0.956], p = 0.022; animal: IRR 0.896, 95% CI [0.821, 0.978], p = 0.014) in early psychosis patients, using negative binomial regression models as appropriate and as adjusted as above (**Fig. 3a, b**).

**Figure 3:**
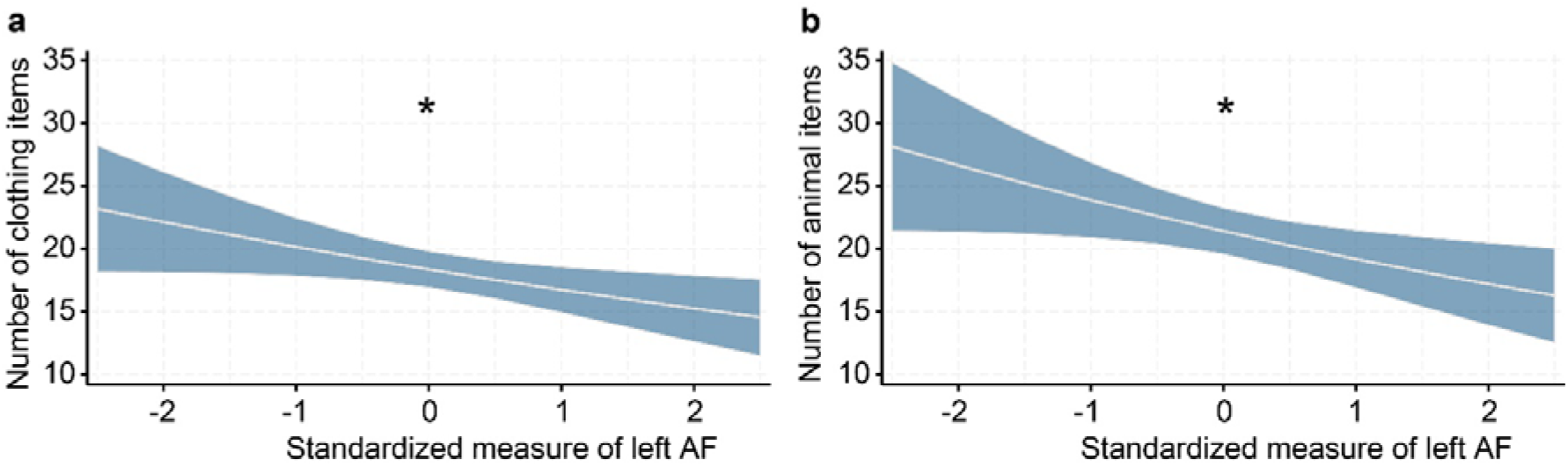
Association between white matter integrity of the arcuate fasciculus and verbal fluency. (**a**) In individuals with early psychosis, fractional anisotropy of the left arcuate fasciculus was negatively associated with the number of clothing items named within one minute. (**b**) A similar negative association was observed for animal items. Standardized measure: z-transformed values of left AF. Shown are predictive margins based on estimates from negative binomial regression models reported above. Shaded areas represent 95% confidence intervals. *P* < 0.05.

No significant correlations between IFOF, UF or ILF and semantic fluency were found in patients with psychosis. Furthermore, in this group, the majority of differences in the associations of fractional anisotropy measures (IFOF, UF, ILF, and AF) with semantic fluency were not significant. Exceptions were AF vs. ILF in relation to clothing and animal naming, where significant differences were found (chi2(1) = 5.80, p = 0.016 and chi2(1) = 5.35, p = 0.021 respectively), as established with seemingly unrelated estimation adjusted as above. No significant group differences were found regarding the corticospinal tract.

## Discussion

Our study provides novel insights into the neural and behavioral aspects of semantic processing in early psychosis. By separating semantic representation from semantic control and linking these dimensions to DTI measures of language stream components, our findings advance the understanding of semantic processing in psychosis. Importantly, this study employed the meticulous procedure of manual fiber tractography on DTI data to reliably assess the integrity of ventral and dorsal pathways.

Individuals with early psychosis showed specific impairments in semantic control, such as reduced verbal fluency and longer response times in a semantic association task compared to individuals with schizotypy. Adjusting for processing speed revealed that these deficits extend beyond general cognitive impairments, highlighting a distinct disruption in semantic control. Our findings of impaired verbal fluency and prolonged response latencies in schizophrenia are consistent with previous reports in the literature ^44–46^. However, we found no significant differences in semantic representation between the groups, suggesting that a primary challenge in early psychosis may lie in handling and manipulating semantic information ^47,48^. In our DTI analysis, we observed reduced mean diffusivity for the left IFOF and left UF in individuals with early psychosis compared to individuals with schizotypy. This finding was unexpected and contrasts with prior studies reporting increased mean diffusivity in ventral language stream components of psychosis patients ^13,49^. However, it aligns with recent evidence suggesting that white matter abnormalities in ventral language stream components show high variability in psychosis patients ^50^. Lower mean diffusivity, as observed in this study, may reflect greater fiber density or axonal integrity, potentially driven by compensatory neurodevelopmental mechanisms during the early stages of psychosis. In contrast, reductions in fiber density, possibly linked to excitotoxic processes ^51^, might become more prominent in later stages of the disorder ^52^. These findings underscore the heterogeneity of psychosis and emphasize the need to account for stage-specific and individual differences when interpreting microstructural white matter alterations. Interestingly, fractional anisotropy in the left AF, a key component of the dorsal language stream, was negatively correlated with verbal fluency in the early psychosis group. This association suggests that lower fractional anisotropy, typically indicative of reduced structural integrity, may mitigate dysregulation of the AF in early psychosis. We did not find significant correlations between ventral language stream DTI measures and semantic control, which may be attributable to the limited sample size. Interestingly, the implication of the left AF in semantic control in our early psychosis cohort suggests that semantic processing in psychosis may not be restricted to the ventral stream ^53^. Our findings indicate that, within the context of psychosis, a strict anatomical separation between phonological processes (dorsal pathway) and conceptually driven language production (ventral pathway) may not exist in such a clear-cut form. This interpretation is consistent with the WEAVER++/ARC model ^54^, which posits that both phonological and semantic processes can be mediated through the AF and therefore do not exhibit a complete anatomical dissociation, also in psychosis.

These findings have important implications for understanding the neural basis of language dysfunction in psychosis. By disentangling semantic representation from semantic control, we highlight the importance of semantic control deficits in early psychosis and their potential association with dorsal stream pathways. Future research should explore the functional dynamics of these pathways using multimodal neuroimaging approaches, such as combining functional MRI with DTI, to better understand the interplay between structural and functional alterations in language networks. Additionally, the temporal dynamics of white matter changes across different stages of psychosis, and their relationship with excitotoxic and neuroinflammatory processes, require further exploration.

## Conclusion

This study advances the field by splitting semantic processing into distinct components and linking these components to specific white matter pathways. Our findings extend traditional models of semantic organization in psychosis, suggesting that semantic control may be organized not only in ventral but also in dorsal language stream components.

## Data Availability

All data produced in the present study are available upon reasonable request to the authors.

## Acknowledgements

This research was supported by grants from the Swiss National Science Foundation (Grant No. 501100001711–191938, to N.D.), the Brain & Behaviour Research Foundation (Grant No. 28997, to W.S.), and the OPO Foundation (Grant No. 2020-0075, to W.S., N.D., P.B. and P.H.). We gratefully acknowledge their financial assistance, which made this work possible.

## Supplementary Information

**Supplementary Table 1:** Behavioral Data.

|  | Low<br>schizotypy (*n* =<br>40) | High<br>schizotypy (*n* =<br>40) | Patient<br>(*n* = 40) |
| --- | --- | --- | --- |
| DO80 |  |  |  |
| Right answer (%) | 92.53 | 93.06 | 87.63 |
| Semantic paraphasia (%) | 1.06 | 0.81 | 1.69 |
| Phonematic paraphasia (%) | 0.19 | 0.09 | 0.72 |
| Neologism (%) | □ | 0.06 | 0.09 |
| Visual error (%) | 0.56 | 0.38 | 0.72 |
| Visuo-semantic error (%) | 4.94 | 4.44 | 6.56 |
| Semantic-phonematic error (%) | □ | □ | 0.03 |
| Visuo-phonematic error (%) | □ | □ | □ |
| Visuo-semantic-phonematic error (%) | □ | □ | □ |
| Did not recognize (%) | 0.22 | 0.19 | 0.78 |
| No answer/word-finding difficulty (%) | 0.16 | 0.28 | 0.47 |
| Description of use/context (%) | 0.03 | 0.13 | 0.28 |
| Other error (%) | 0.13 | 0.31 | 0.41 |
| Morphematic neologism (%) | 0.19 | 0.25 | 0.63 |
| MWT-B Total, *M* (*SD*) | 24.68 (3.7) | 24.85 (4.16) | 22.8 (4.11) |
| CCT |  |  |  |
| Correct items [%], *M* (*SD*) | 0.84 (0.08) | 0.84 (0.07) | 0.8 (0.1) |
| Reaction time [s], *M* (*SD*) | 3.01 (0.82) | 3.47 (1.09) | 4.89 (2.36) |
| Semantic Verbal Fluency |  |  |  |
| Animals: No. of correct items, *M* (*SD*) | 25.98 (6.15) | 24.65 (7.47) | 21.25 (6.16) |
| Clothes: No. of correct items, *M* (*SD*) | 25.1 (5.68) | 22.38 (5.99) | 18.35 (4.82) |
| Food: No. of correct items, *M* (*SD*) | 29.2 (6.7) | 25.63 (5.41) | 22.65 (6.71) |

**Supplementary Figure 1:**
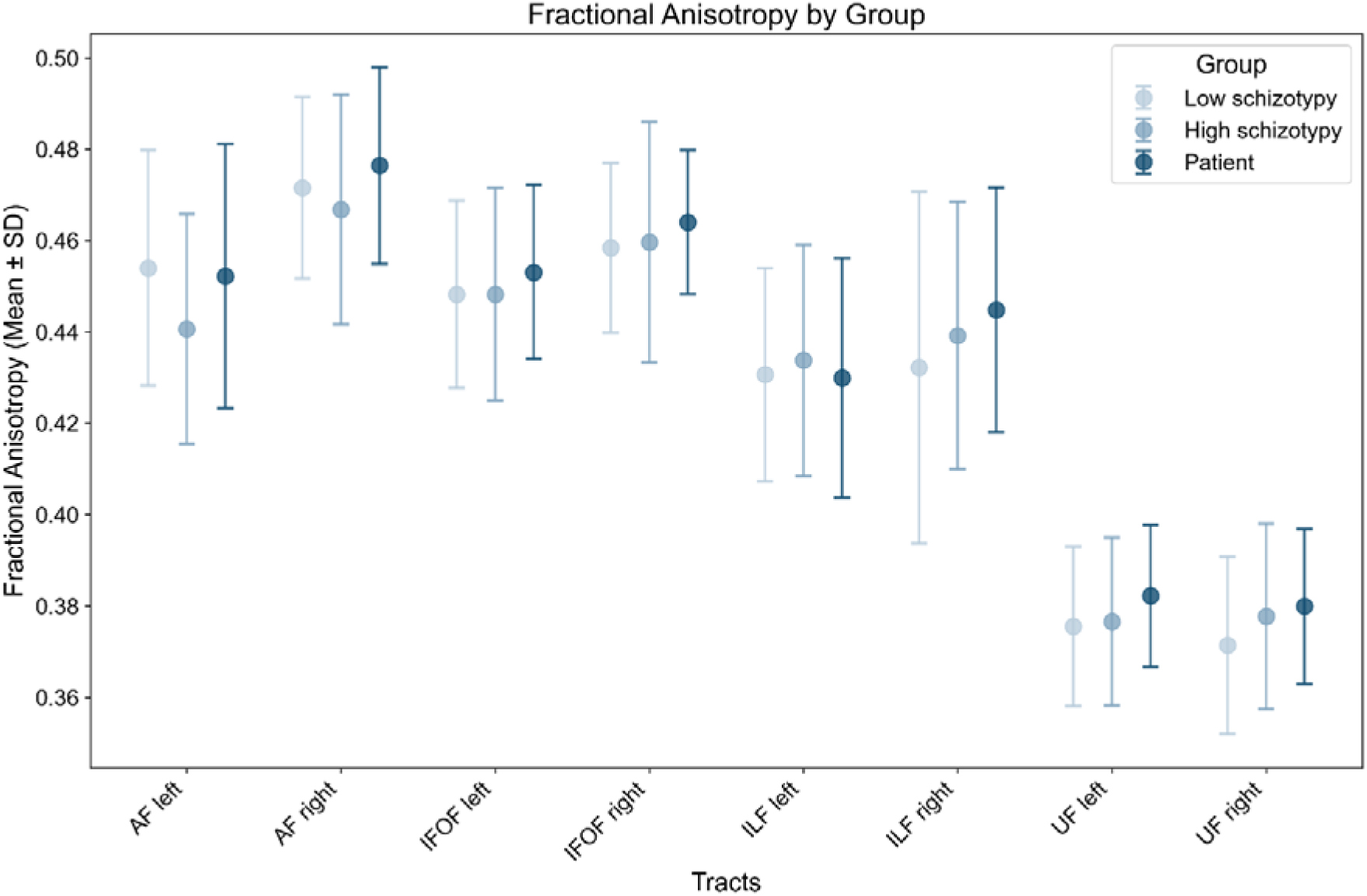
Diffusion tensor imaging metrics across language tracts. Fractional anisotropy (mean ± 1 standard deviation (SD)) is plotted for ventral and dorsal pathways, shown separately for low-schizotypy controls (n = 40), high-schizotypy controls (n = 40), and early-psychosis patients (n = 40).

**Supplementary Figure 2:**
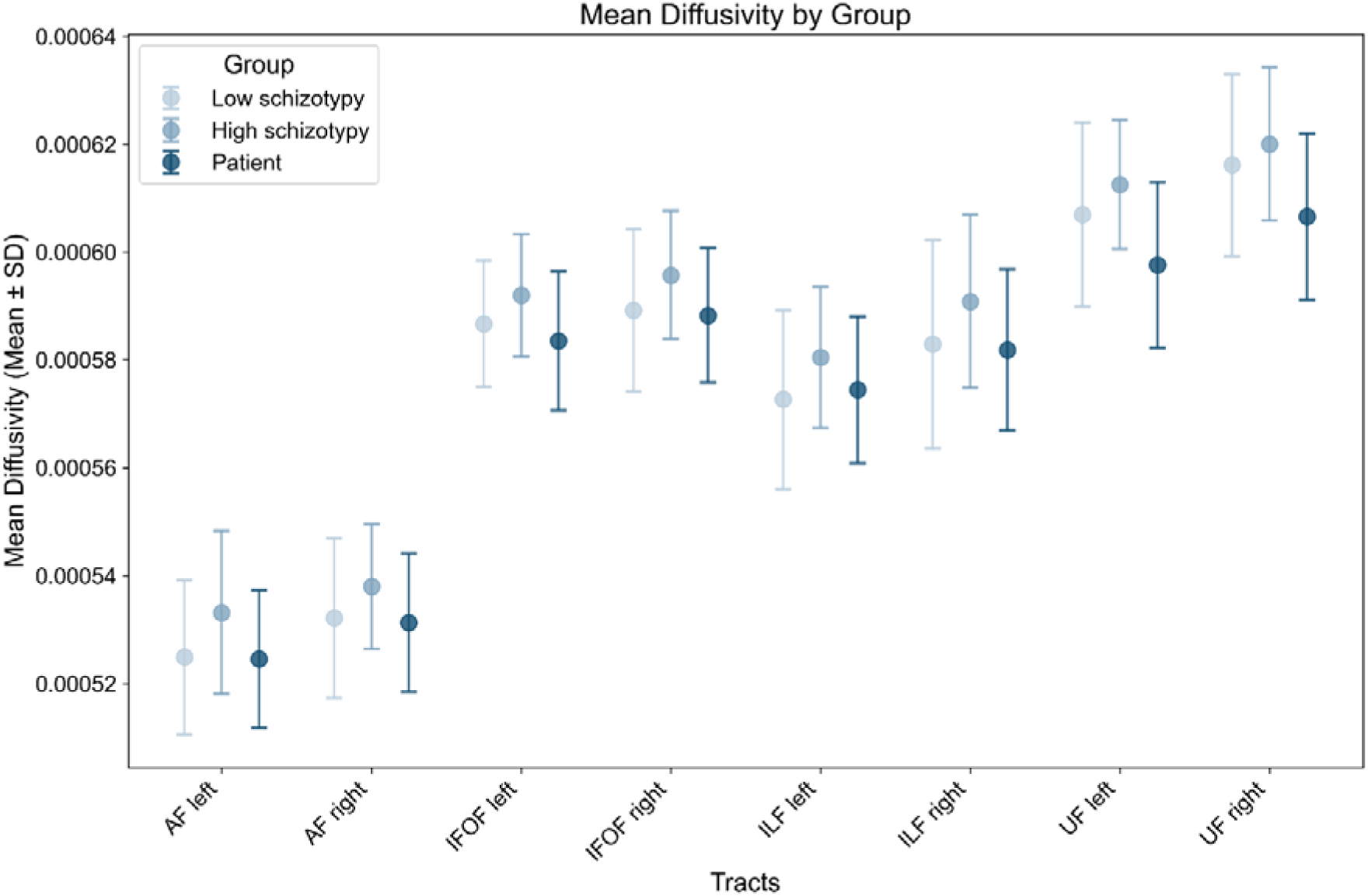
Diffusion tensor imaging metrics across language tracts. Mean diffusivity (mean ± 1 standard deviation (SD)) is plotted for ventral and dorsal pathways, shown separately for low-schizotypy controls (n = 40), high-schizotypy controls (n = 40), and early-psychosis patients (n = 40).

